# Accurate Skin Lesion Classification Using Multimodal Learning on the HAM10000 and ISIC 2017 Datasets

**DOI:** 10.1101/2024.05.30.24308213

**Authors:** Abdulmateen Adebiyi, Nader Abdalnabi, Emily Hoffman Smith, Jesse Hirner, Eduardo J. Simoes, Mirna Becevic, Praveen Rao

**Affiliations:** Department of Electrical Engineering and Computer Science, University of Missouri, Columbia, United States; MU Institute for Data Science and Informatics, University of Missouri, Columbia, United States; Department of Dermatology, University of Missouri, Columbia, United States; Department of Biomedical Informatics, Biostatistics and Medical Epidemiology, University of Missouri, Columbia, United States; Department of Dermatology, Saint Louis University, Saint Louis, United States

**Keywords:** Multimodal Deep Learning, Skin Lesion Classification, Primary Care Physician, Skin Cancer Screening

## Abstract

**Background:** Our aim is to demonstrate that multimodal deep learning can enhance the accuracy of classifying skin lesions using both images and textual descriptions (e.g., demographics, clinical information) compared to a classifier that learn only on images.

**Methods:** We used the HAM10000 and ISIC 2017 datasets in our study containing 10,000 and 2,750 skin lesion images, respectively. We combined the images with patients’ data (e.g., sex, age, lesion location) for training and evaluating a multimodal deep learning classification model. The dataset was split into 70% for training the model, 20% for the validation set, and 10% for the testing set. We compared the multimodal model’s performance to well-known deep learning models that only use images for classification.

**Results:** We used accuracy and area under the curve (AUC) receiver operating characteristic (ROC) as the metrics to compare the models’ performance. Our multimodal model outperformed the competitors and achieved the best results. Our model’s accuracy and AUCROC was 0.9411 and 0.9426, respectively, on HAM10000. On ISIC 2017, our model’s accuracy and AUCROC was 0.7971 and 0.8253, respectively.

**Conclusion:** Our study showed that a multimodal deep learning model can outperform traditional deep learning models for skin lesion classification on the HAM10000 and ISIC 2017 datasets. Our approach can enable primary care clinicians to screen for skin cancer in patients (residing in areas lacking access to expert dermatologists) with higher accuracy and reliability.

## Background

Skin cancer is the most common type of cancer diagnosed worldwide^1^. It is estimated that approximately 9,500 people in the United States (US) are diagnosed with skin cancer each year^1^. It is predicted that around 20% of people in the US will develop skin cancer in their lifetime^2^. The two most common skin cancer types are basal cell cancer and squamous cell cancer, while melanoma is the third most common skin cancer^2^. However, melanoma has the highest mortality, with a long-term survival of less than 10%, despite recent decline in mortality attributed to improved screening and better treatment such as immunotherapy^3^.

There are geographical differences in the incidence and mortality of skin cancer^4^. The incidence and mortality of melanoma is higher for individuals living in rural and underserved areas, than their urban counterparts^4^. Many factors may contribute to this geographical disparity in melanoma incidence and death, including increased ultraviolet radiation exposure and lower adoption of sun protection strategies in rural compared to urban residents^5^. In addition, barriers to health care access and availability contributes to late detection and effective treatment of skin cancers. The lack of adequate number and distribution of dermatologists contribute to late detection of melanoma. Patients face considerable wait times ranging from 33.9 to 73.4 days to consult with a dermatologist regarding changing moles. Interestingly, even when medical care is offered at no cost, a significant number of patients decline it if the travel distance for their appointment exceeds 20 miles^6^.

This situation is accentuated in isolated rural areas. Tele-dermatology has proven effective in mitigating geographic isolation, and various studies have affirmed its diagnostic and treatment accuracy and reliability through telemedicine. Nonetheless, numerous obstacles hinder its widespread adoption and implementation. Alongside privacy and liability concerns, dermatologists identify the absence of a consistent reimbursement system as the primary impediment for both *store-and-forward* (i.e., clinical information is used asynchronously to patient encounter) and *live-interactive* (i.e., synchronous) tele-dermatology^7^. Karavan et al. discovered that 40% of patients diagnosed with melanoma in traditional clinics resided in areas where tele-dermatology services were underutilized^8^.

Primary care clinicians based (PCCs) in the community often serve as the initial point of contact for patients and may assume a crucial role in offering screening and early diagnosis, particularly for individuals lacking sufficient access to dermatologists. PCCs, while essential, face limitations compared to dermatologists in terms of early detection and have identified insufficient training during medical school and residency as hindrances to effective skin screening^7,9^. In primary care settings, the rapid and accurate diagnosis of skin conditions is of paramount importance for patient care^9^. However, distinguishing between various skin lesions can be a challenging task, and misdiagnosis can lead to serious consequences^11^.

Prior work showed the effectiveness of state-of-the-art deep learning models for skin lesion classification (into malignant and benign classes) using dermascopy images^12^. With growing interest in multimodal deep learning models^13^, it is now possible to combine skin lesion images and textual data (e.g., lesion location, patient age) for model training and inference. A multimodal deep learning model learns by integrating multiple modalities of input data (e.g., image and text) and enabling it to be more precise for downstream tasks (e.g., classification) compared to conventional deep learning models that learn on one modality (e.g., image). In this work, *we investigate whether multimodal models can improve the accuracy of skin cancer diagnosis compared to models that only use images*. Our approach well-known publicly available datasets comprising a diverse range of dermatological images^10,14^ including basal cell cancer and melanoma.

## Methods

### Dataset

In this study, we used two publicly available datasets. The first is the well-known HAM10000 dataset^14^, which is a large dermatoscopic image collection of common pigmented skin lesions. This dataset was collected over a duration of 20 years from two countries, namely, Austria and Australia. The images were categorized into 7 classes, namely, Actinic Keratoses (AKIEC), Basal Cell Carcinoma (BCC), Benign Keratosis (BKL), Dermatofibroma (DF), Melanocytic Nevi (NV), Melanoma (MEL), and Vascular Skin Lesion (VASC). Table *1* shows examples of skin lesion images from each class along with the number of images per class. The HAM10000 dataset has the Age, Sex and Lesion Location metadata. The second is the well-known ISIC 2017 Dataset^10^. It was collected from 10+ international dermatology clinics. This dataset contains 2,750 dermatoscopic images and has the Age and Sex metadata; the images were categorized into 3 classes, namely MEL, Seborrheic Keratosis (SK), and Benign Nevus (BN). (Description of HAM10000/ISIC 2017 Table in the supplementary materials shows the brief description of the variables in the HAM10000/ISIC 2017 datasets.)

**Table 1:**
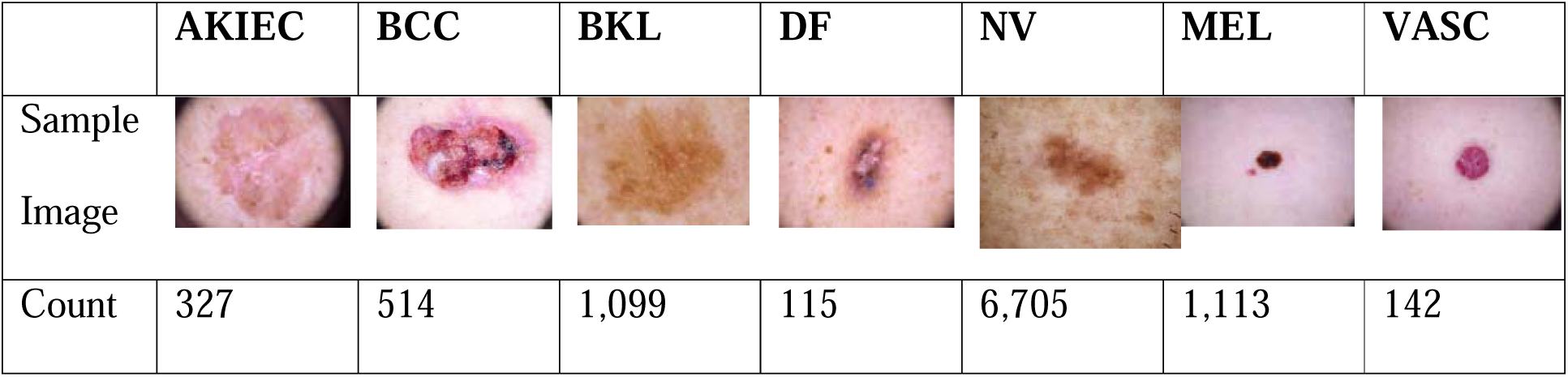
Examples of skin lesions in HAM10000.

|  | **AKIEC** | **BCC** | **BKL** | **DF** | **NV** | **MEL** | **VASC** |
| --- | --- | --- | --- | --- | --- | --- | --- |
| Sample Image |  |  |  |  |  |  |  |
| Count | 327 | 514 | 1,099 | 115 | 6,705 | 1,113 | 142 |

### Deep Learning Models

In recent years, deep learning has gained a lot of traction in image understanding, image classification, language translation, speech recognition, and natural language processing. Convolutional neural networks (CNNs) have shown excellent performance in large-scale image classification and object detection competitions such as the ImageNet Large Scale Visual Recognition Challenge (ILSVRC)^15^. We briefly discuss a few popular CNNs and the multimodal model that we used in our work.

CNNs that are deep with many layers are prone to overfitting and consume a lot of computational resources. **Inception-V3** was introduced by Szegedy et. al. in 2014^16^ to solve these problems by using sparsely connected architectures. It uses the inception module that applies multiple convolutions (e.g., 1×1 convolution, 3×3 convolution, 5×5 convolution) and a maximum pooling layer. The outputs are concatenated to create the input for the next stage.

He et. al. introduced the deep residual neural network (**ResNet**) architecture^17^ that uses skip connections between layers to solve the vanishing gradient problem. Residual blocks reduced the total parameters by allowing the gradient to flow directly through the skipped connections backward from later layers to the initial filter. In our work, we used ResNet50 with 50 layers.

Huang et. al. introduced the **DenseNet**^18^ architecture where every layer in the model is connected to every other layer in a feed-forward manner. DenseNet was proposed to solve the problem of vanishing gradient while being computationally efficient. It promotes feature reuse resulting in a more compact model. In our work, we used DenseNet121 with 121 layers.

**EfficientNet** was developed by Tan et. al. to achieve high accuracy and computational efficiency^19^. It balances the network width, depth, and resolution to build a competitive yet computationally efficient CNN model. The compound coefficient technique is used to scale up models in an effective way in EfficientNet. We used EfficientB0 in our work.

Junnan Li et. al. introduced the **ALBEF** model^20^ that learns the joint representation of image and text data. The model combines the Vision Transformer^21^ (ViT-b/16) as the image encoder and BERT^22^ as the text encoder. We used the joint text-image encoder to encode both the text and images, and then added a linear fully connected layer to it based on the number of output classes. The Image encoder were initialized with weights pre-trained on ImageNet-1k^23^. The input image was encoded into a sequence of embeddings.

For ALBEF, we set hyperparameters such as the patch size to 16, embedding dimension to 768, depth to 12, number of attention heads to 12, multilayer perceptron (MLP) ratio to 4, and query-key-value bias to True. The forward pass of the model combines the representation from the Image Encoder Outputs (Image Embeddings and Image Attentions) and the text encoder outputs (Text Embeddings and the Text Attention Mask). We used early fusion in combing the two representations. We performed the fusion in the text encoder with the aid of an attention-based alignment, where the text encoder jointly processes text and image features. We then added a fully connected layer for the classification.

### Data Pre-processing

Each original image in HAM10000 was of size 600×450 pixels. The images in ISIC 2017 varied from 542×718 pixels to 2,848×4,288 pixels. As different deep learning models used different image sizes as input, we had to resize the original images. For ResNet50 and DenseNet121, the images were resized to 224×224 pixels. For Inception-V3, the images were resized to 299×299 pixels. For EfficientNetB0, the images were resized to 224×224 pixels. Finally, for ALBEF, the images were resized to 256×256 pixels.

### Skin Lesion Classification Approach

We split each dataset into three sets randomly: the training set (containing 70% of the images), the validation set (containing 20% of the images), and the testing set (containing 10% of the images). The training and validation sets were used for the training phase. We then used the testing set to evaluate the trained models. The test set in HAM10000 had the following number of images for each class: AKIEC: 38, BCC: 49, BKL: 110, DF: 11, MEL: 109, NV: 667, and VASC: 18. The test set in ISIC 2017 had the following number of images for each class: MEL: 55, SK: 194, and BN: 27.

Figure 1 shows the overall approach for skin lesion classification. Our system was implemented in Python using PyTorch^24^, CUDA, Numpy^25^, and OpenCV^26^ libraries. We used existing implementations of the ALBEF model for our multimodal lesion classification^27^. The models were trained and tested on a Dell Precision server with an Intel Xeon processor,96 GB RAM, 2 TB disk storage, and two NVIDIA Quadro RTX4000 (8GB) graphics processing units (GPUs).

**Figure 1:**
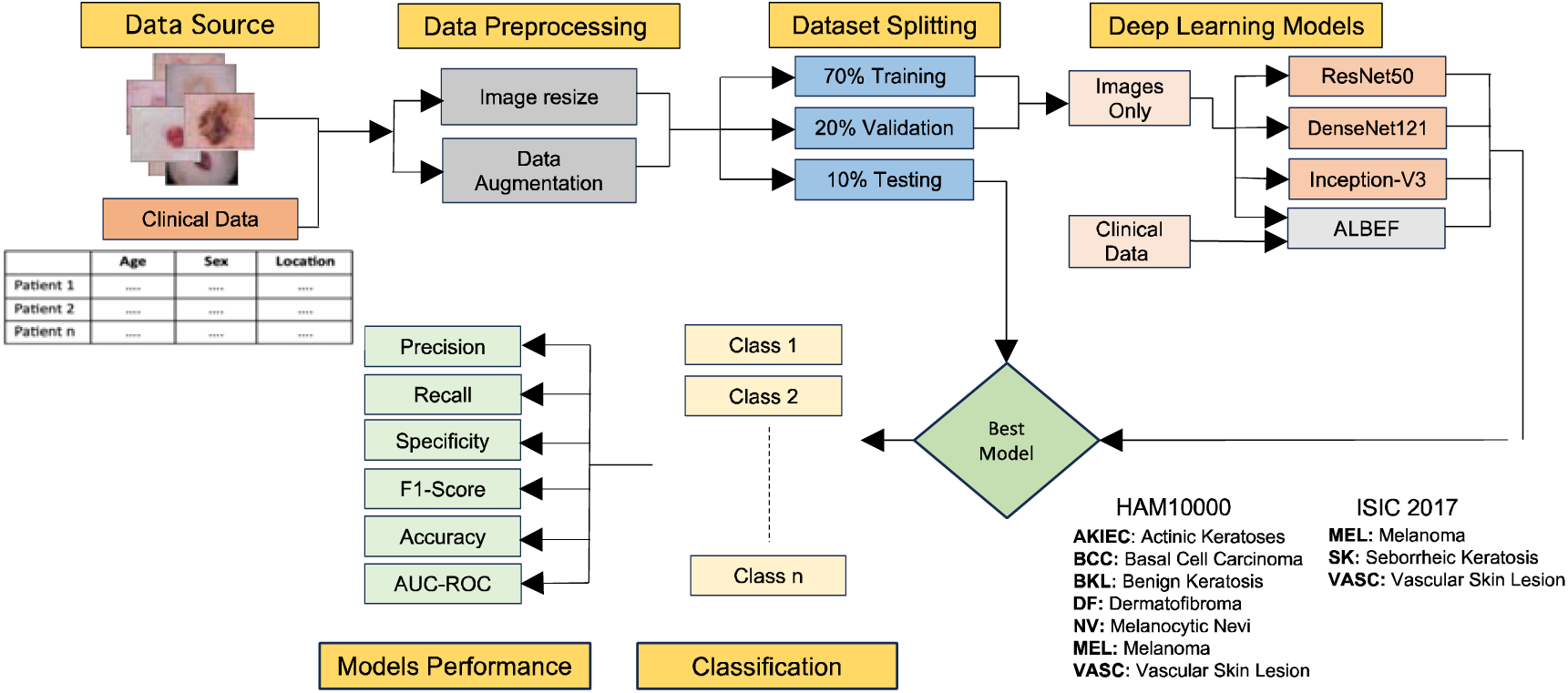
Skin lesion classification approach

### Model Training Settings

The Inception-V3, ResNet50, DenseNet121, EfficientNetB0, and ALBEF models were trained with the same hyperparameters: (a) batch size of 16, (b) 200 epochs, and (c) learning rate of 1e-4. Also, each model used the Adam optimizer^28^ and binary cross entropy loss function. The best model based on the highest validation accuracy was saved and used for classification on the test set.

Data augmentation is a widely used technique by deep learning models to prevent overfitting (during training) and enhance the generalization of the models on unseen data. It is useful when a dataset is imbalanced. For the Inception-V3, ResNet50, EfficientNetB0, and DenseNet121 model, we applied data augmentation during training by performing the transformations (a) random horizontal flip, (b) random vertical flip, (c) random rotation, and (d) color jitter. We trained on 70% of the dataset and picked our best model by using 20% of the dataset as the validation set.

In our ALBEF model, we resized the image to 256 pixels. For data augmentation, we performed the transformations (a) random resized crop and (b) random horizontal flip. Our other hyperparameters were weight decay of 0.01 and Epsilon (eps) of 1e-4. Similar to other models, we trained on 70% of the dataset and picked our best model by using 20% of the dataset as the validation set.

In addition to resizing the images during data pre-processing, we also normalized the images. We obtained the mean and standard deviation of the images for the normalization step. Normalization ensures that all features are scaled uniformly, which prevents any single feature from disproportionately dominating the skin lesion classification. This process enhances model stability and accuracy.

## Results

In this section, we present the performance of InceptionV3, ResNet50, DenseNet121, EfficientNetB0, and the Multimodal fusion (ALBEF) on the HAM10000 and ISIC 2017 datasets. For the ALBEF model, we considered two different settings: 1) Use the images with the associated text (e.g., age, sex, lesion location) for training the dataset. 2) Use only the images and passing blank text for training on the datasets. We did this to show the effect of adding demographics/clinical information on the overall performance of the model.

### Performance Metrics

Next, we briefly describe the performance metrics that we used for evaluating our different models. The classification models aim to classify the ISIC 2017 test set into 3 classes and the HAM10000 test set into 7 classes. We used true positives (Tp), true negatives (Tn), false positives (Fp), and false negatives (Fn) in the computation of our different performance metrics. Below are the brief definitions of Tp, Fp, Fn, and Tn. Tp indicates the total number of lesion images that were predicted correctly in the positive class. Tn denotes the total number of lesion images that are in the negative class and are classified correctly as the negative class. Fn denotes the total number of lesion images that were in the positive class but were predicted incorrectly as the negative class. Fp denotes the total number of lesion images that are in the negative class but were predicted incorrectly as the positive class.

The different metrics used in our evaluation are listed below:

1. Precision (P): Tp/(Tp+Fp)
2. Sensitivity/Recall (R): Tp/(Tp+Fn)
3. Specificity: Tn/(Tn+Fp)
4. Accuracy: (Tp+Tn)/(Tp+Tn+Fp+Fn)
5. F1-score: 2×P×R/(P+R)

Table 2 shows the performance achieved by InceptionV3, ResNet50, DenseNet121, EfficientNetB0 and ALBEF on the HAM10000 and ISIC 2017 datasets. ALBEF (Text and Images) outperformed all other models in terms of accuracy and AUCROC on both the datasets. This demonstrates that by fusing images and textual data (e.g., age/sex, age/sex/lesion location) via multimodal deep learning can enhance the classification accuracy of deep learning models.

**Table 2:** Performance comparison of different models (best result is shown in bold)

| Dataset | Models | Accuracy | Precision | Sensitivity | Specificity | F1-Score | AUCROC |
| --- | --- | --- | --- | --- | --- | --- | --- |
|  | InceptionV3 | 0.7899 | 0.7058 | **0.7087** | **0.8377** | 0.7042 | 0.7732 |
| ISIC 2017 | ResNet50 | 0.7717 | 0.6636 | 0.6571 | 0.8252 | 0.6567 | 0.7411 |
|  | DenseNet121 | 0.7935 | 0.7521 | 0.6805 | 0.8225 | **0.7132** | 0.7126 |
|  | EfficientNet | 0.7500 | 0.6472 | 0.6744 | 0.8095 | 0.6480 | 0.7515 |
|  | ALBEF (Only Images) | 0.7681 | 0.6705 | 0.6550 | 0.8081 | 0.6520 | 0.7316 |
|  | ALBEF (Text and Images) | **0.7971** | **0.7522** | 0.6475 | 0.8114 | 0.6805 | **0.8253** |
| HAM10000 | InceptionV3 | 0.8653 | 0.8202 | 0.7608 | 0.9570 | 0.7838 | 0.8589 |
|  | ResNet50 | 0.8503 | 0.7815 | 0.7129 | 0.9646 | 0.7292 | 0.8388 |
|  | DenseNet121 | 0.8862 | 0.8505 | 0.8249 | 0.9685 | 0.8350 | 0.8967 |
|  | EfficientNet | 0.9022 | 0.8784 | 0.8664 | 0.9717 | 0.8714 | 0.9191 |
|  | ALBEF (Only Images) | 0.9132 | 0.8959 | 0.8524 | 0.9748 | 0.8693 | 0.9136 |
|  | ALBEF (Text and Images) | **0.9411** | **0.9073** | **0.9019** | **0.9833** | **0.9033** | **0.9426** |

### Confusion Matrices and AUCROC Plots

We also computed the confusion matrices for the ALBEF model on the two datasets. These matrices are shown in Figures 2 and 3. We also constructed the AUCROC plots of different models on the two datasets. These plots are shown in Figures 4 and 5.

**Figure 2:**
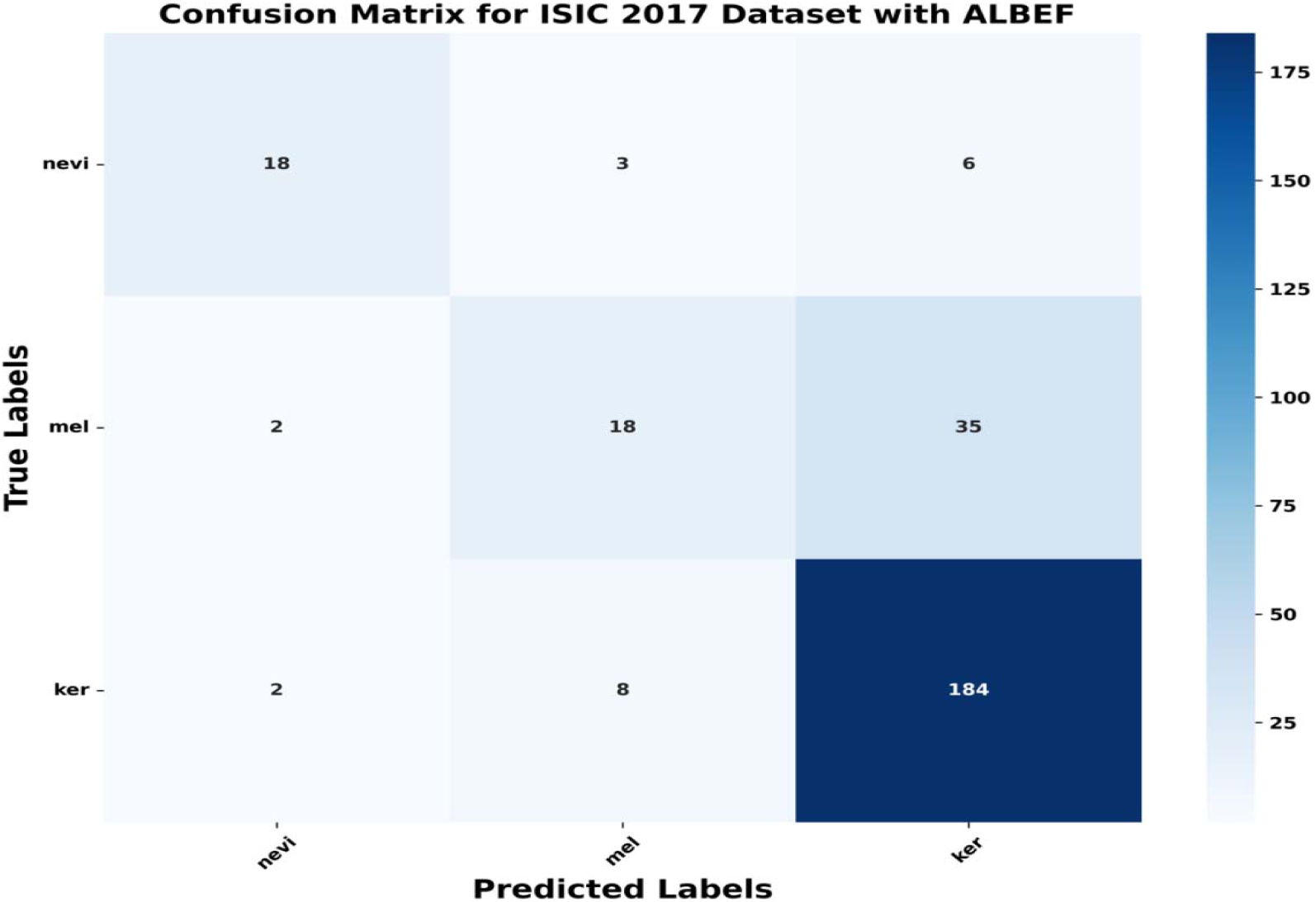
Confusion matrix for the ISIC 2017 dataset with the ALBEF Model

**Figure 3:**
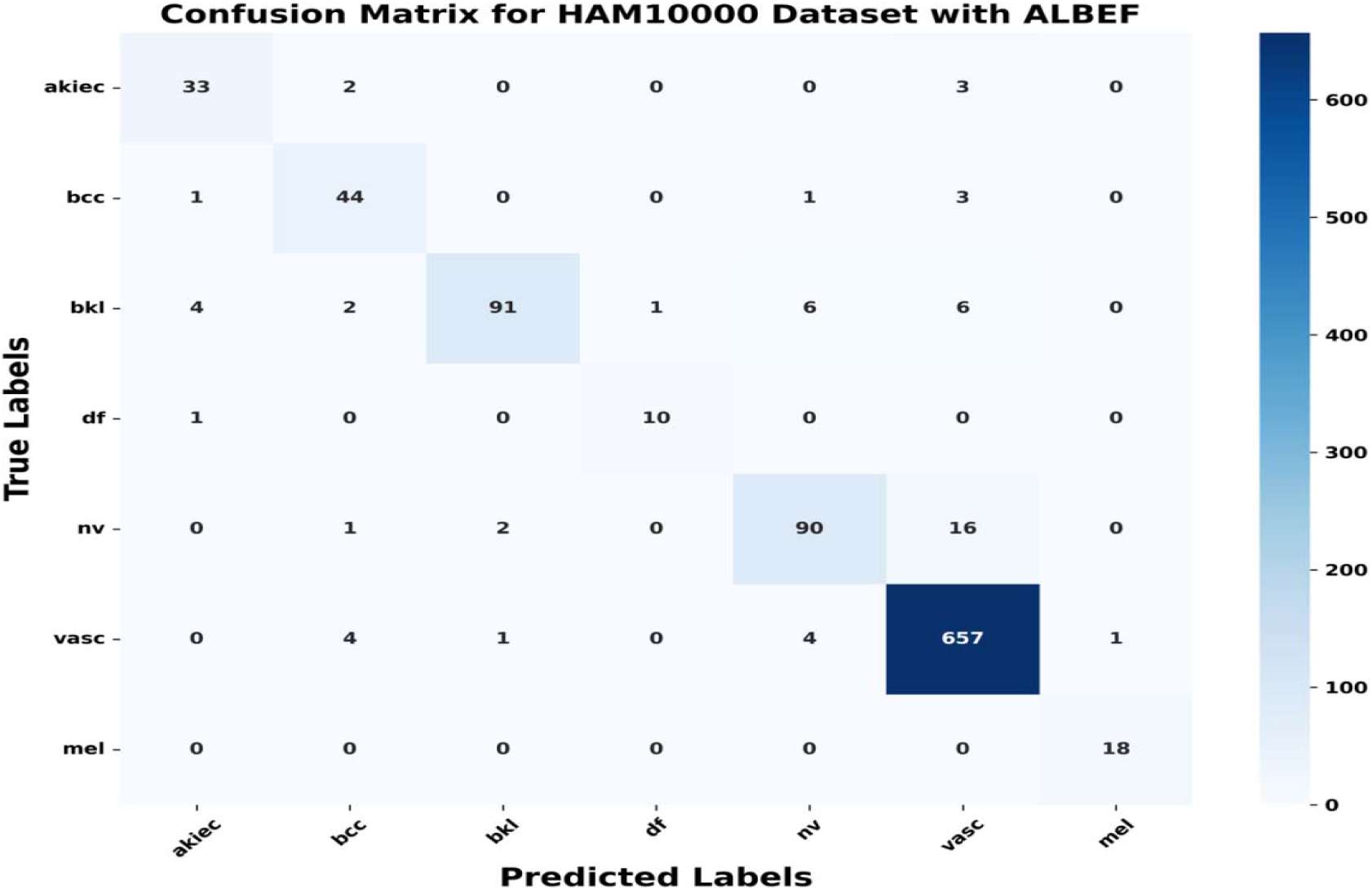
Confusion matrix for the HAM10000 dataset with the ALBEF Model

**Figure 4:**
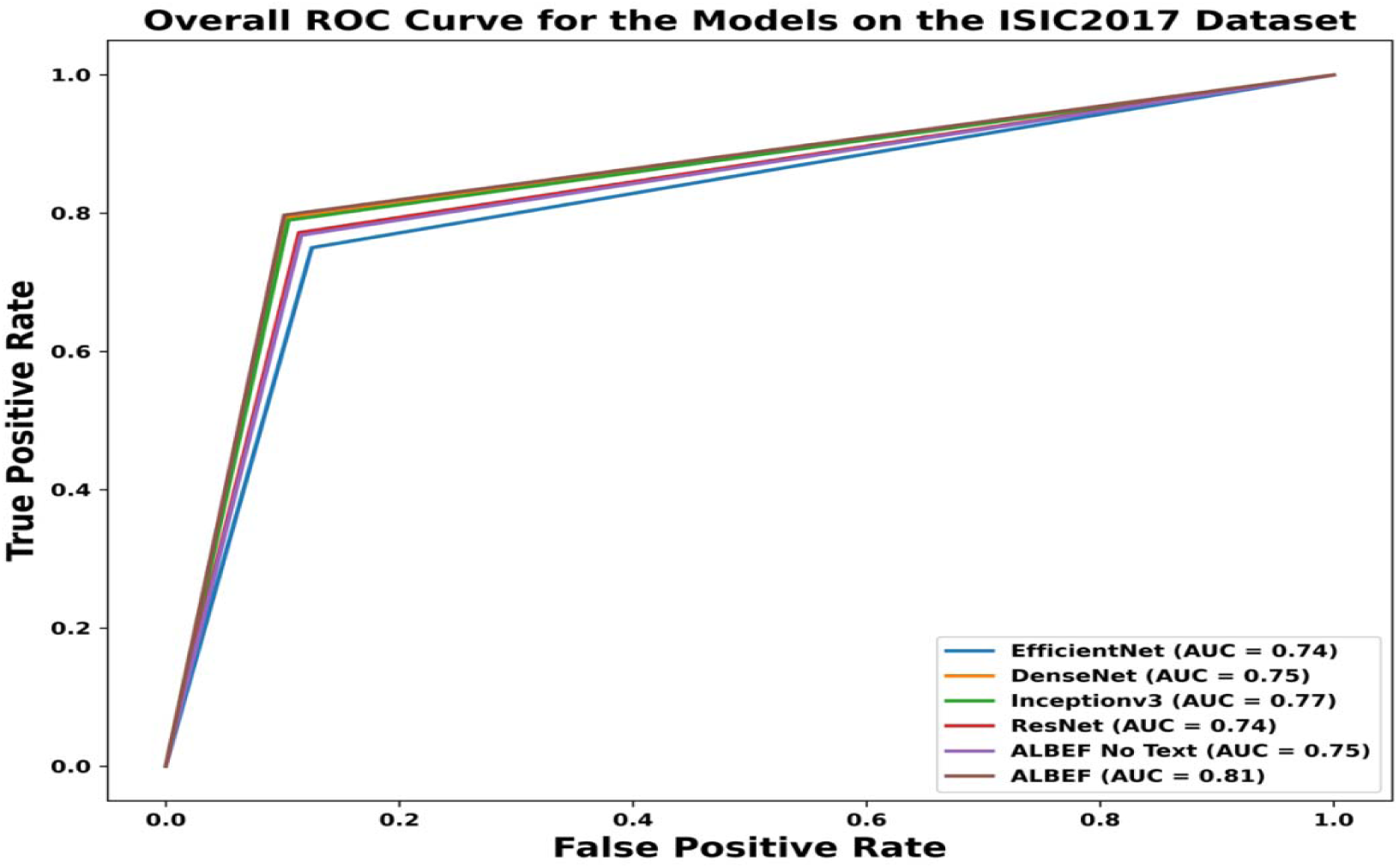
AUCROC plot for the ISIC 2017 dataset

**Figure 5:**
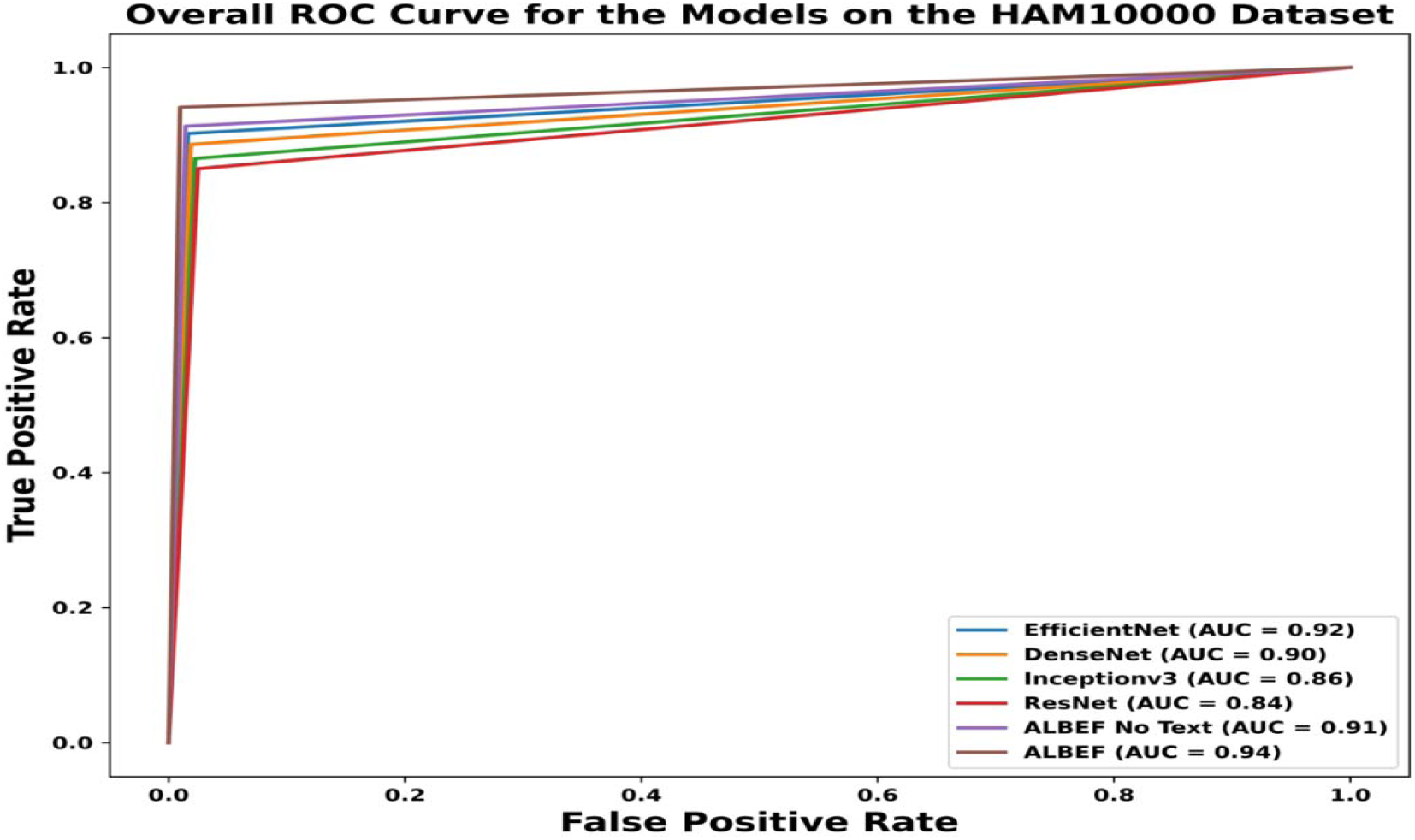
AUCROC plot for the HAM10000 dataset

### Ablation Study

An ablation study is a technique in deep learning that is used to determine the impact of different components/features in a model. This study enabled us to understand the contribution of the different features in the ALBEF models. Table 3 below shows the results of the ablation study on ISIC 2017 and HAM10000 datasets. Clearly, the multimodal model achieved the best classification performance when all the available textual features were used along with the lesion images during training/testing.

**Table 3:** Ablation study results on ALBEF using different textual features (best result in bold)

| Dataset | Model | Accuracy | AUCROC |
| --- | --- | --- | --- |
| ISIC 2017 | ALBEF (Image w/ Age) | 0.7645 | 0.7451 |
|  | ALBEF (Image w/ Sex) | 0.7754 | 0.6856 |
|  | ALBEF (Image w/ Age, Sex) | **0.7971** | **0.8253** |
| HAM10000 | ALBEF (Image w/ Age) | 0.9112 | 0.9064 |
|  | ALBEF (Image w/ Sex) | 0.9152 | 0.9197 |
|  | ALBEF (Image w/ Localization) | 0.9002 | 0.8992 |
|  | ALBEF (Image w/ Age, Sex, Localization) | **0.9411** | **0.9426** |

Finally, we compared the classification accuracy of our multimodal model (i.e., ALBEF) with other models published in the literature on the HAM10000 dataset. Our model achieved higher accuracy compared to other models reported in Table 4. A caveat is that these models were trained with different hyperparameters.

**Table 4:** Comparison with prior work on HAM10000.

| Prior Work | Model(s) | Accuracy |
| --- | --- | --- |
| Prasad et al.<sup>29</sup> | MobileNet | 0.8310 |
| Laghari et al.<sup>30</sup> | CNN-transfer learning | 0.8790 |
| Dhivya et al.<sup>31</sup> | CNN | 0.9000 |
| Luo et al.<sup>32</sup> | RegNetY-320 | 0.9100 |
| Diwan et al.<sup>33</sup> | InceptionResNetV2 + ResNetXt101 | 0.9283 |
| **Our Model** | **ALBEF** | **0.9411** |

## Discussion

Multimodal deep learning provides the ability to combine information from multiple models. Information from different sources like audio, text, image, and video assists in implementing more complex models that improve the performance of many applications^34^.

The ALBEF model enables us to fuse visual and clinical information, providing a holistic view of the skin lesions^20^. This multimodal approach has the potential to significantly enhance the diagnostic capabilities of primary care physicians and nurse practitioners. ALBEF has been used in a wide array of domains such as hateful detection, image retrieval etc^20^. ALBEF can learn joint representations, which has made it very useful in image retrieval tasks such as in text queries, text-based on images e-commerce applications, etc. It has also been applied in natural language processing for generating text captions for images^20^.

In this study, we employed ALBEF to classify the skin lesions in the HAM10000 and ISIC 2017 datasets. In addition to the lesion images, ALBEF was trained on the lesion images as well as textual metadata such as Age, Sex, and Localization. The AUCROC score and accuracy of classification of ALBEF was the best compared to other models. For HAM10000, ALBEF (lesion images w/ Age, Sex and Localization) achieved an accuracy of 0.9411 and AUCROC score of 0.9426. For ISIC 2017, ALBEF (lesion images w/ Age and Sex) achieved an accuracy of 0.7971 and AUCROC score of 0.8253. Furthermore, based on the ablation study, we observed that the performance of ALBEF improved as more textual features were considered. Hence, our multimodal approach is deemed applicable in a primary care setting where high accuracy and robustness is desired.

Recently, Adebiyi et al applied three different models (InceptionV3, ResNet50 and DenseNet121) for their skin lesion classification^12^. They collected 770 de-identified dermoscopy images from the University of Missouri (MU) Healthcare. After applying a hair removal algorithm on the lesion images, they observed that DenseNet121 achieved the best accuracy of 0.8052 and an AUCROC score of 0.81 in their experiments. They did not use a multimodal approach.

Alam et al achieved an accuracy of 0.85 on the HAM10000 dataset by using the Inception-V3 model^32^. Akter et al achieved an accuracy of 0.82 on the HAM10000 dataset using the ResNet50 model^35^. Our ALBEF model, on the other hand, achieved an accuracy of 0.9411.

Tschandi et al applied multimodal learning for skin lesion classification^36^. They employed two ResNet50 CNNs followed by a late fusion technique to combine the features. Their result showed that combining the dermoscopic with macroscopic images and the metadata improves the performance. Multimodal machine learning has been applied in Knee Osteoarthritis progression prediction from plain radiographs and clinical data by Tiulpin et al^37^. They utilized the raw radiographic data, clinical examination results, and previous medical history of the patient. They were able to achieve an AUCROC score of 0.79.

Harahap and Zulkifli utilized InceptionResNet-v2 for skin lesion image classification, achieving 0.8756 accuracy on the HAM10000 dataset^38^. They employed transfer learning and TensorFlow Lite for efficient model deployment in a web application, enhancing accessibility for users seeking skin disease diagnosis. Anisi et al. trained their model by using 2,000 and 8,695 dermoscopic images from ISIC 2017 and ISIC 2018 respectively^39^. They found out that YOLO^40^ achieved a better accuracy score and is faster compared to the pre-existing classifiers. Finally, Useni et al.^41^ employed a self-supervised artificial intelligence (AI) approach for skin lesion classification. It achieved an expert-level accuracy in distinguishing ‘ugly ducklings’ from average-looking skin lesions in wide-field images.

Deep learning models often lack interpretability. They are considered black box in nature; therefore, it may be difficult to understand how the classifications are achieved which is important for clinical decision making. SHAP (SHapley Additive exPlanations) was proposed recently to address this issue^42^. SHAP assigns a Shapley value to the pixels in an image classification task. It highlights which part of the lesion had the most impact in the deep learning model decision. It helps to know the key features a model relies on for its prediction. It also helps in understanding how a deep learning model differentiates and interprets various classes in an image classification task. The SHAP results on model explainability are provided in the supplementary document for DenseNet121. (Note that we were unable to generate SHAP plots for ALBEF.)

Deep learning-based applications in the healthcare sector have shown significant and promising solutions for timely and accurate diagnosis. Studies have demonstrated the potential of deep learning to “transform medical diagnostics”, with enhanced diagnostic accuracy of algorithms using medical imaging such as X-ray, CT scans or MRIs for various diseases and conditions.^43,44^ For skin diseases, real-life deployment of deep learning models can help bridge specialty access to care gaps by guiding non-primary clinicians, who are typically the first to encounter skin conditions, with clinical decision-making. Developing and validating deep learning models require inclusion of collaborating dermatologists to identify and elaborate on key clinical features likely to influence clinical outcomes, such as age (melanoma prevalence and types of skin conditions often vary by age), sex (autoimmune diseases may affect sexes differently due to hormonal influences and immune system differences), or lesion localization (some skin conditions may be site-specific (rosacea primarily affects the central face or seborrheic dermatitis that affects areas rich in sebaceus glands, such as the scalp) ^45^.

Our study has some limitations. We only used three kinds of metadata (Age, Sex, and Lesion Location) with the images in our multimodal classification. Having additional metadata in the dataset can further improve the classification performance. Another issue is the imbalanced test sets for HAM10000 and ISIC 2017. Some classes had small number of images. While we used data augmentation during training, weighted classification metrics, which consider the number of test examples in each class, can be computed for tackling the data imbalance issue.

## Conclusion

In our study, we showed that multimodal model specifically the ALBEF model can outperform the traditional deep learning models such as Inception-V3, ResNet50, DenseNet121, EfficientNetB0 in the early detection of skin lesions on the HAM10000 and ISIC 2017 datasets. This can assist primary care physicians and nurse practitioners to screen for skin cancer in patients (residing in areas lacking access to expert dermatologists) with higher accuracy and reliability. Our study also showed that the fusion of images and textual data (e.g., age/sex, age/sex/lesion location) via multimodal deep learning can enhance the classification accuracy of deep learning models.

## Funding Statement

This work was supported by a **T**ranslational **R**esearch **I**nforming **U**seful and **M**eaningful **P**recision **H**ealth (TRIUMPH) grant from the University of Missouri-Columbia.

## Competing Interests

The authors have no competing interests to declare.

## Contributions

All authors contributed to conception and design of the work, drafting or critical revision of the manuscript and final approval of the manuscript. All authors agree to be accountable for all aspects of the work.

## Ethics declarations

### Ethics approval and consent to participate

Not applicable

### Consent for publication

Not applicable

### Competing interests

The authors declare no competing interests

## Data Availability

The HAM10000 is available at https://doi.org/10.7910/DVN/DBW86T; the ISIC 2017 dataset is available at https://challenge.isic-archive.com/data/#2017.

## Notes

### Competing Interest Statement

The authors have declared no competing interest.

### Funding Statement

This project was funded by the Translational Research Informing Useful and Meaningful Precision Health (TRIUMPH) grant from the University of Missouri-Columbia.

### Author Declarations

This study used only publicly available dataset. The HAM10000 and ISIC2017 datasets.

### Summary of Updates

This is the uppdated version

## References

1. World Health Organization, International Agency for Research on Cancer. Skin cancer. Retrieved, from https://www.iarc.who.int/cancer-type/skin-cancer

2. American Academy of Dermatology. *Available from* https://www.aad.org/media/stats-skin-cancer

3. National Cancer Institute Surveillance, Epidemiology, and End Results (SEER) Program. Melanoma of the skin - Cancer stat facts. from https://seer.cancer.gov/statfacts/html/melan.html

4. Blake KD, Moss JL, Gaysynsky A, Srinivasan S, Croyle RT. Making the Case for Investment in Rural Cancer Control: An Analysis of Rural Cancer Incidence, Mortality, and Funding Trends. Cancer Epidemiology, Biomarkers & Prevention: A Publication of the American Association for Cancer Research, Cosponsored by the American Society of Preventive Oncology

5. Kalia S, Kwong Y k. k., Haiducu M l., Lui H. Comparison of Sun Protection Behaviour Among Urban and Rural Health Regions in Canada. Journal of the European Academy of Dermatology and Venereology. 2013;27(11):1452–4.

6. Pala P, Bergler-Czop BS, Gwiżdż JM. Teledermatology: Idea, Benefits and Risks of Modern Age – a Systematic Review Based on Melanoma. Journal of Telemedicine and Telecare. 2020 Apr;37(2):159–67.

7. Jones OT, Jurascheck LC, van Melle MA, Hickman S, Burrows NP, Hall PN, et al. Dermoscopy for Melanoma Detection and Triage in Primary Care: A Systematic Review. BMJ open. 2019 Aug 20;9(8):e027529.

8. Karavan M, Compton N, Knezevich S, Raugi G, Kodama S, Taylor L, et al. Teledermatology in the Diagnosis of Melanoma. Journal of Telemedicine and Telecare. 2014 Jan;20(1):18–23.

9. Brown AE, Najmi M, Duke T, Grabell DA, Koshelev MV, Nelson KC. Skin Cancer Education Interventions for Primary Care Providers: A Scoping Review. Journal of General Internal Medicine. 2022 Jul;37(9):2267–79.

10. Codella NCF, Gutman D, Celebi ME, Helba B, Marchetti MA, Dusza SW, et al. Skin Lesion Analysis Toward Melanoma Detection: A Challenge at the 2017 International Symposium on Biomedical Imaging (ISBI), Hosted by the International Skin Imaging Collaboration (ISIC) arXiv; 2017 https://arxiv.org/abs/1710.05006

11. Li H, Pan Y, Zhao J, Zhang L. Skin Disease Diagnosis with Deep Learning: A Review. Neurocomputing. 2021 Nov 13;464:364–93.

12. Adebiyi A, Rao P, Hirner J, Anokhin A, Hoffman Smith E, Simoes E, and Becevic M. Comparison of Three Deep Learning Models in Accurate Classification of 770 Dermoscopy Skin Lesion Images. In AMIA 2024 Informatics Summit, 8 pages, Boston, 2024.

13. Huang Y, Du C, Xue Z, Chen X, Zhao H, Huang L. What Makes Multi-Modal Learning Better than Single (Provably). In: Advances in Neural Information Processing Systems. Curran Associates, Inc.; 2021. p. 10944–56.

14. Tschandl P, Rosendahl C, Kittler H. The HAM10000 dataset, a large collection of multi-source dermatoscopic images of common pigmented skin lesions. Sci Data. 2018 Aug 14;5(1):180161.

15. Russakovsky O, Deng J, Su H, Krause J, Satheesh S, Ma S, et al. ImageNet Large Scale Visual Recognition Challenge 2015 http://arxiv.org/abs/1409.0575

16. Szegedy C, Liu W, Jia Y, Sermanet P, Reed S, Anguelov D, et al. Going deeper with convolutions. In: 2015 IEEE Conference on Computer Vision and Pattern Recognition (CVPR). 2015. p. 1–9.

17. He K, Zhang X, Ren S, Sun J. Deep Residual Learning for Image Recognition. In: 2016 IEEE Conference on Computer Vision and Pattern Recognition (CVPR). 2016. p. 770–8.

18. Huang G, Liu Z, Van Der Maaten L, Weinberger KQ. Densely Connected Convolutional Networks. In: 2017 IEEE Conference on Computer Vision and Pattern Recognition (CVPR). 2017. p. 2261–9.

19. Tan M, Le QV. EfficientNet: Rethinking Model Scaling for Convolutional Neural Networks. arXiv; 2020.

20. Li J, Selvaraju RR, Gotmare AD, Joty S, Xiong C, Hoi S. Align Before Fuse: Vision and Language Representation Learning with Momentum Distillation. arXiv; 2021.

21. Dosovitskiy A, Beyer L, Kolesnikov A, Weissenborn D, Zhai X, Unterthiner T, et al. An Image is Worth 16×16 Words: Transformers for Image Recognition at Scale. In: International Conference on Learning Representations 2021.

22. Devlin J, Chang MW, Lee K, Toutanova K. BERT: Pre-training of Deep Bidirectional Transformers for Language Understanding. In: Burstein J, Doran C, Solorio T, editors. Proceedings of the 2019 Conference of the North American Chapter of the Association for Computational Linguistics: Human Language Technologies, Volume 1 (Long and Short Papers) [Internet]. Minneapolis, Minnesota: Association for Computational Linguistics; 2019. p. 4171–86.

23. Deng J, Dong W, Socher R, Li LJ, Li K, Fei-Fei L. Imagenet: A Large-Scale Hierarchical Image Database. In: 2009 IEEE Conference on Computer Vision and Pattern Recognition. 2009. p. 248–55.

24. Paszke A, Gross S, Massa F, Lerer A, Bradbury J, Chanan G, et al. Pytorch: An Imperative Style, High-Performance Deep Learning Library. In: Advances in Neural Information Processing Systems. Curran Associates, Inc.; 2019.

25. Harris CR, Millman KJ, van der Walt SJ, Gommers R, Virtanen P, Cournapeau D, et al. Array programming with NumPy. Nature. 2020 Sep;585(7825):357–62.

26. Bradski G. The Opencv Library. Vol. 25, Dr. Dobb’s J. Softw. Tools. 2000.

27. Multimodal-Learning-Hands-on-Tutorial/multimodal_training.ipynb at main · dsaidgovsg/multimodal-learning-hands-on tutorial/blob/main/multimodal_training.ipynb

28. Kingma DP, Ba J. Adam: A Method for Stochastic Optimization 2017

29. Chaturvedi SS, Gupta K, Prasad PS. Skin Lesion Analyser: An Efficient Seven-Way Multi-class Skin Cancer Classification Using MobileNet. In: Hassanien AE, Bhatnagar R, Darwish A, editors. Advanced Machine Learning Technologies and Applications. Singapore: Springer; 2021. p. 165–76.

30. Ali K, Shaikh ZA, Khan AA, Laghari AA. Multiclass skin cancer classification using EfficientNets – a first step towards preventing skin cancer. Neuroscience Informatics. 2022 Dec 1;2(4):100034.

31. G D, K L, M M, K N, C T. Skin Cancer Detection Using Multi Class CNN Algorithm. In: 2023 9th International Conference on Advanced Computing and Communication Systems (ICACCS). 2023. p. 1353–6.

32. Alam TM, Shaukat K, Khan WA, Hameed IA, Almuqren LA, Raza MA, et al. An Efficient Deep Learning-Based Skin Cancer Classifier for an Imbalanced Dataset. Diagnostics. 2022 Sep;12(9):2115.

33. Chaturvedi SS, Tembhurne JV, Diwan T. A Multi-Class Skin Cancer Classification Using Deep Convolutional Neural Networks. Multimedia Tools and Applications. 2020 Oct 1;79(39):28477–98.

34. Pei X, Zuo K, Li Y, Pang Z. A Review of the Application of Multi-modal Deep Learning in Medicine: Bibliometrics and Future Directions. International Journal of Computational Intelligence Systems. 2023 Mar 29;16(1):44.

35. Akter MS, Shahriar H, Sweta Sneha. Multi-Class Skin Cancer Classification Architecture Based on Deep Convolutional Neural Network. In: 2022 IEEE International Conference on Big Data (Big Data). 2022. p. 5404–13.

36. Yap J, Yolland W, Tschandl P. Multimodal Skin Lesion Classification Using Deep Learning. Experimental Dermatology. 2018 Nov;27(11):1261–7.

37. Tiulpin A, Klein S, Bierma-Zeinstra SMA, Thevenot J, Rahtu E, Meurs J van, et al. Multimodal Machine Learning-based Knee Osteoarthritis Progression Prediction from Plain Radiographs and Clinical Data. Scientific Reports. 2019 Dec 27;9(1):20038.

38. Harahap NI, Zulkifli FY. Web Application Development Skin Lesion Classification Using Transfer Learning InceptionResNet-v2. IJECBE. 2023 Dec 30;1(2):65–75.

39. Singh SK, Abolghasemi V, Anisi MH. Fuzzy Logic with Deep Learning for Detection of Skin Cancer. Applied Sciences. 2023 Jan;13(15):8927.

40. Redmon J, Divvala S, Girshick R, Farhadi A. You Only Look Once: Unified, Real-Time Object Detection.2016

41. Useini V, Tanadini-Lang S, Lohmeyer Q, Meboldt M, Andratschke N, Braun RP, et al. Automatized self-supervised learning for skin lesion screening. Sci Rep. 2024 Jun 3;14(1):12697.

42. Lundberg SM, Lee SI. A Unified Approach to Interpreting Model Predictions. In: Advances in Neural Information Processing Systems. Curran Associates, Inc.; 2017.

43. Song Y, Zheng S, Li L, Zhang X, Zhang X, Huang Z, et al. Deep Learning Enables Accurate Diagnosis of Novel Coronavirus (COVID-19) With CT Images. IEEE/ACM Transactions on Computational Biology and Bioinformatics. 2021 Nov;18(6):2775–80.

44. Zhen S hui, Cheng M, Tao Y bo, Wang Y fan, Juengpanich S, Jiang Z yu, et al. Deep Learning for Accurate Diagnosis of Liver Tumor Based on Magnetic Resonance Imaging and Clinical Data. Frontiers in Oncology. 10.

45. Aggarwal R, Sounderajah V, Martin G, Ting DSW, Karthikesalingam A, King D, et al. Diagnostic Accuracy of Deep Learning in Medical Imaging: A Systematic Review and Meta-Analysis. npj Digital Medicine. 2021 Apr 7;4(1):1–23.

